# Diastolic Dysfunction Predicts Cardiorespiratory Fitness and Clinical Outcomes in Hypertrophic Cardiomyopathy

**DOI:** 10.64898/2026.10.23.26366319

**Authors:** Mark E. Pepin, Jack O’Sullivan, Diona Gjermeni, Everton J. Santana, Bryan Mosher, Sanjay Malunjkar, Jason V. Tso, Euan A. Ashley, Eldrin Lewis, Jeffrey W. Christle, Jonathan N. Myers, Victoria Parikh, Masataka Kawana, Matthew T. Wheeler, Francois Haddad

## Abstract

**Background:** Echocardiographic indices of diastolic dysfunction are routinely obtained in hypertrophic cardiomyopathy (HCM), but their relationships with cardiorespiratory fitness and long-term clinical outcomes remain poorly defined.

**Objectives:** This study therefore sought to determine the extent to which E/e’, left atrial volume index (LAVi), and peak tricuspid regurgitation velocity (TRV_max_) relate to peak oxygen uptake, ventilatory efficiency, their longitudinal trajectories, and clinical outcomes in HCM.

**Methods:** We retrospectively studied patients with HCM who completed cardiopulmonary exercise testing within 7 days of resting echocardiography. Each diastolic index was related to percent-predicted peak oxygen uptake (V̇O_2_), ventilatory efficiency (V̇E/V̇CO_2_ slope), their serial trajectories, and a composite of heart failure hospitalization, transplantation, or death.

**Results:** Among 441 patients with HCM (mean age 49 ± 16 years; 66% men), each SD higher E/e’, LAVi, and TRV_max_ was independently associated with lower peak V̇O_2_ (−6.0, −3.6, and −6.3 percentage points of predicted) and higher V̇E/V̇CO_2_ slope (+0.9, +1.0, and +2.5; all *P* ≤ 0.045), nonlinearly for E/e’ with peak V̇O_2_ and LAVi with V̇E/V̇CO_2_ slope. On serial testing, only LAVi predicted progressively worsening V̇E/V̇CO_2_ slope (+0.4/yr/SD; 95% CI: 0.13-0.68; *P* = 0.005). Over a median of 4.8 years, 118 of 436 patients (27.1%) reached the composite endpoint, for which LAVi was the strongest echocardiographic predictor (adjusted HR: 1.68/SD; 95% CI: 1.40-2.03).

**Conclusions:** In HCM, resting diastolic indices were associated with cardiorespiratory fitness, and higher LAVi was independently associated with a steeper rise in V̇E/V̇CO_2_ slope and heart failure events. Resting diastolic phenotyping may complement exercise testing.

## Introduction

Hypertrophic cardiomyopathy (HCM) is the most common inherited cardiovascular condition and is associated with a chronic, progressive decline in cardiorespiratory fitness that frequently culminates in heart failure with preserved ejection fraction (HFpEF).^1,2^ Formalized exercise capacity testing is often used to gauge clinical status and disease prognosis, such that randomized therapeutic trials have employed cardiopulmonary exercise testing (CPET) as a quantitative measure of disease severity.^3,4^ Resting echocardiography, by contrast, is obtained in nearly every patient, yet how its measures of diastolic function relate to exercise capacity and its trajectory remains uncertain.

Exercise limitation in HCM is multifactorial. Diastolic abnormalities are among the earliest features of HCM and can be detected in genotype-positive individuals with impaired early relaxation before hypertrophy develops.^5–9^ Nevertheless, the relationship between individual diastolic indices and CPET measures remains inadequately characterized.^10,11^

The echocardiographic assessment of left ventricular diastolic dysfunction (LVDD) in HCM relies on Doppler-derived surrogates of mean left atrial pressure and LV relaxation.^12^ Many standard measurements are confounded by dynamic loading conditions and coincident valvular abnormalities, resulting in only modest correlations with invasive hemodynamics.^13,14^ Earlier guideline-based algorithmic strategies of diastolic classification compound these issues, yielding a high proportion of indeterminate cases and failing to capture the continuous spectrum of LVDD.^15^

Society recommendations have since adopted tailored approaches for LVDD characterization in HCM to lessen the reliance on preload-dependent measurements.^14,16^ Specifically, updated guidance now recommends the use of average E/e’ (>14), Ar-A (≥30 ms, the difference between pulmonary venous atrial reversal duration and transmitral A-wave), left atrial maximum volume index (LAVi >34 mL/m^2^), and peak tricuspid regurgitation velocity (TRV_max_ >2.8 m/s) that together exhibit a robust correlation with invasive LV filling pressures.^17–20^ Successive multicenter analyses support this approach in HFpEF, showing improved sensitivity relative to the previous framework.^21^ Larger cohort analyses, including the Atherosclerosis Risk in Communities study, also support age-stratified reference ranges and establish the prognostic relevance of diastolic function for incident heart failure complications and mortality.^22^ However, these analyses did not address functional capacity or clinical trajectory, nor did they include individuals with HCM.

This study therefore characterizes the relationship between diastolic dysfunction and cardiorespiratory fitness in a large retrospective cohort of HCM. Specifically, we describe the continuous relationship of individual echocardiographic LVDD indices with peak V̇O_2_ and V̇E/V̇CO_2_ slope using multivariable models with restricted cubic splines. We also incorporate serial exercise testing and long-term clinical outcomes to determine whether LVDD is associated with fitness trajectory and adverse clinical events.

## Methods

### Ethical approval

The study protocol was reviewed and approved by the Stanford University Institutional Review Board (protocol 25673, The Cardioshare Project). Patient data were de-identified prior to analysis, and results are reported in aggregate to preserve individual anonymity. ***Study population and patient selection.*** A retrospective cohort of patients with HCM who had undergone CPET at Stanford University Hospital between October 2006 and August 2021 was analyzed, before the commercial availability of cardiac myosin inhibitors, wherein HCM was defined in accordance with AHA/ACC guidelines as a maximum septal or posterior wall thickness (IVSd or LVPWd) ≥1.5 cm, an adjudicated apical HCM phenotype, or a wall thickness ≥1.3 cm in patients harboring a known pathogenic HCM variant.^2^ To minimize the confounding effects of cardiorespiratory comorbidities, we excluded patients with obstructive coronary artery disease, chronic obstructive pulmonary disease, or interstitial lung disease. All remaining CPETs were aligned with the closest resting echocardiogram within 7 days of testing. Using this approach, we identified 441 subjects with recorded age, sex, body mass index (BMI), and a valid CPET measurement (**Figure 1**). Because complete diastolic assessment was not universally available, indices were analyzed in the patients for whom it was measured. To compare indices within the same patients, a prespecified common cohort of 261 patients in whom both E/e’ and LAVi were measured was also analyzed. Of the 441 patients, E/e’ was measured in 316 and LAVi in 301; accordingly, 40 patients had LAVi without E/e’, and 55 had E/e’ without LAVi. Non-HCM reference subjects (n = 337) were drawn from the same registry after exclusion of HCM, LVEF <50%, wall thickness ≥1.3 cm, or resting LVOT gradient ≥30 mmHg, and were compared with patients with HCM using propensity-score overlap weights for age, sex, and BMI (**Figure S1**).

**Figure 1:**
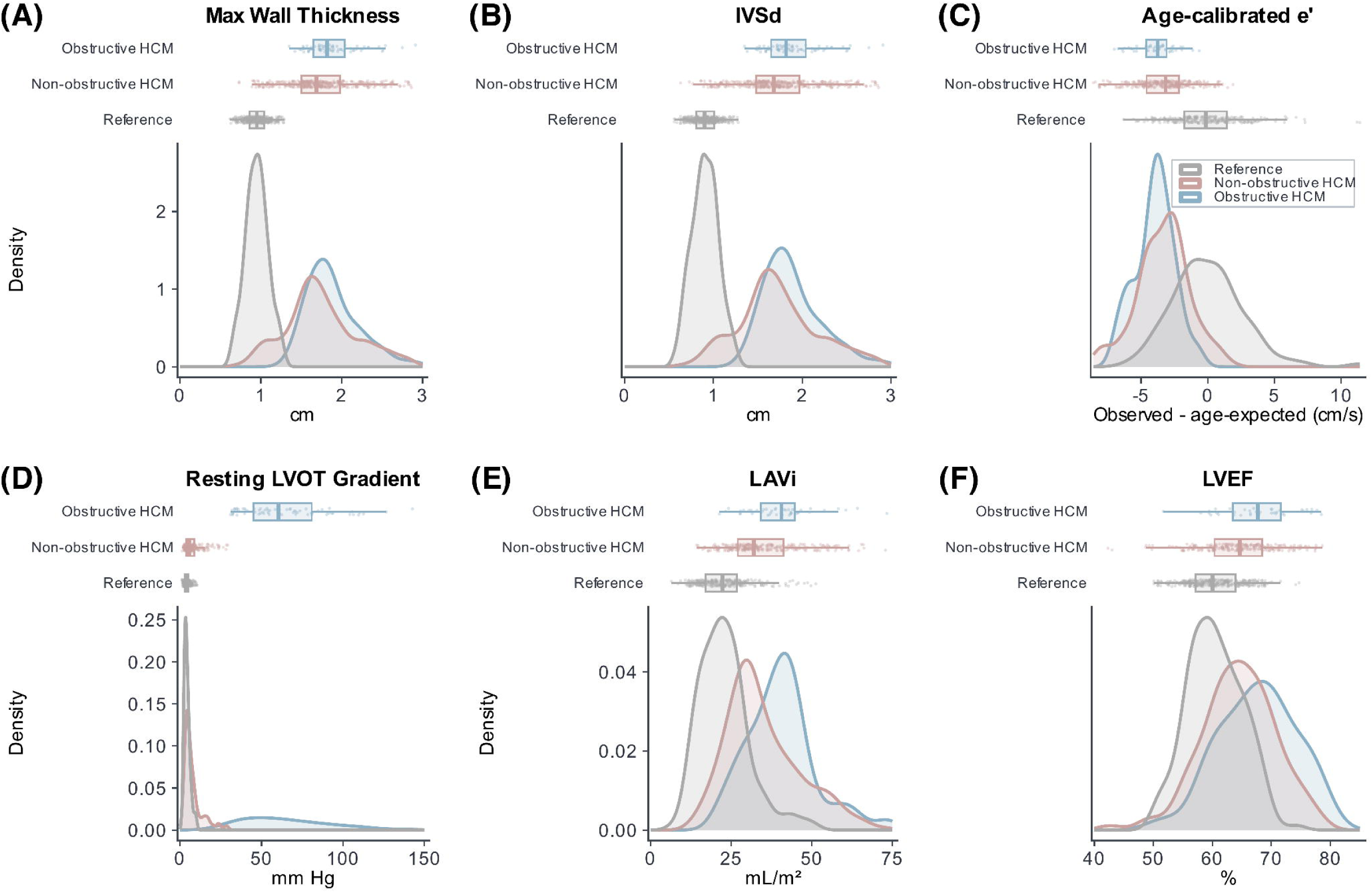
Comparing HCM and non-HCM reference cohorts. The comparison between 441 with HCM and 337 phenotypically screened non-HCM comparators. Densitometric distributions showing **(A)** Maximum wall thickness, **(B)** Interventricular septal thickness in diastole (IVSd), **(C)** Age-calibrated average e’, **(D)** Resting left ventricular outflow tract (LVOT) gradient, **(E)** Left atrial volume index (LAVi), and **(F)** Left ventricular ejection fraction (LVEF). Non-HCM references (gray), non-obstructive HCM (red), and obstructive HCM, defined by resting LVOT gradient ≥30 mm Hg (blue).

### Echocardiography

Resting transthoracic echocardiography was performed using a standardized institutional protocol in accordance with published standards.^23^ Transmitral E- and A-wave velocities were measured by pulsed-wave Doppler, and septal and lateral e’ by tissue Doppler imaging. Diastolic function was characterized using the HCM-specific guidance of the 2025 American Society of Echocardiography (ASE) recommendations,^16^ which specify the following indices with established abnormality thresholds: average E/e’ ratio (>14), left atrial volume index (LAVi >34 mL/m^2^), and peak tricuspid regurgitation velocity (TRV_max_ >2.8 m/s).^15,16^ Average e’ was also compared using age-specific reference limits (<9 cm/s at 20-39 years, <7 cm/s at 40-65 years, and <6.5 cm/s at >65 years).^16^ Septal thickness was measured as the end-diastolic interventricular septal dimension (IVSd) in the parasternal long-axis view.

### Cardiopulmonary exercise testing

Symptom-limited CPET was performed on a treadmill (n = 425) or cycle ergometer (n = 16) with continuous electrocardiography and gas exchange monitoring, as previously reported.^24^ To ensure adequate exertion, we included only tests achieving a respiratory exchange ratio ≥1.0. Peak V̇O_2_ was defined as the highest 20-second average during the final stage and expressed as percent predicted using the 2018 FRIEND reference equation, which incorporates age, sex, weight, height, and exercise mode.^25^ V̇E/V̇CO_2_ slope was derived by linear regression of minute ventilation against carbon dioxide output.

### Cross-sectional association between diastolic indices and cardiorespiratory fitness

To characterize the relationship between LVDD and cardiorespiratory fitness, we modeled each combination of diastolic index and CPET measure using multivariable regression with 4-knot restricted cubic splines, adjusted for age, sex, and BMI. We then quantified the incremental contribution of each index with a partial F-test relative to base covariates alone and tested for departure from linearity by comparing each spline with a linear term (nested F-test). As a sensitivity test, we repeated each model with additional adjustment for LV septal thickness, resting LVOT gradient, beta-blocker and non-dihydropyridine calcium-channel blocker use, and hypertension (**Table S1**). A single breakpoint in each relationship was tested by parametric bootstrap (2,000 replicates), comparing the best-fitting hinge model with the linear model.

### Longitudinal modeling

Repeated CPET measurements were analyzed among patients who underwent 2 or more echo-aligned studies separated by at least 6 months. Studies performed after surgical myectomy were excluded, and follow-up was restricted to 10 years. For each of E/e’, LAVi, and TRV_max_, we modeled percent-predicted peak V̇O_2_ and V̇E/V̇CO_2_ slope in separate linear mixed-effects models with follow-up time, the standardized baseline index, their interaction, age, sex, and BMI as fixed effects, and patient-level random intercepts and slopes (**Table S2**). The time-by-index interaction tested whether baseline diastolic burden modified the subsequent trajectory. As a prespecified exploratory check on the linearity of these trajectories, each model was refit as a generalized additive mixed model (GAMM) with a smooth function of follow-up time and a time-by-index interaction.

### Heart failure outcomes

As a secondary analysis, clinical outcomes were defined from International Classification of Diseases diagnosis and procedure codes in the electronic health record, together with the recorded dates of hospitalization, heart transplantation, and death. The primary outcome end-point was the first of hospitalization for acute heart failure, heart transplantation, or all-cause death after the baseline echo-aligned CPET. Time-to-event analysis was performed from the baseline echo-aligned CPET to the first qualifying event, with censoring at the last documented clinical encounter or January 2025, whichever occurred first. Each index was evaluated in a separate Cox model per SD, adjusted for age, sex, and BMI, and peak V̇O_2_, V̇E/V̇CO_2_ slope, LV septal thickness, and resting LVOT gradient were examined in parallel models (**Table S3**). Proportional hazards were assessed with scaled Schoenfeld residuals (**Table S4**). Event-free survival was compared by log-rank test across E/e’ >14, LAVi >34 mL/m^2^, V̇E/V̇CO_2_ slope >30, and peak V̇O_2_ <80% of predicted. Surgical myectomy was used as a separate secondary end-point, adjusted for resting LVOT gradient.

### Statistical analysis

Continuous variables are reported as mean ± SD or median (IQR) and categorical variables as n (%), using available observations; CPET measures were compared across E/e’–LAVi phenotypes by Kruskal-Wallis test, and event-free survival was estimated by the Kaplan-Meier method. The false discovery rate (FDR) was controlled within each of the cross-sectional, longitudinal, and time-to-event analysis families using the Benjamini-Hochberg method, and adjusted q-values are reported alongside *P*-values. Missing adjustment covariates were retained by modeling unrecorded status as a separate category for binary variables and by pairing the resting LVOT gradient with an indicator of whether it was recorded. Prespecified sensitivity analyses included restriction to adults (≥18 years), the consistency cohorts with 2 or all 3 indices measured, and multiple imputation of missing E/e’ and LAVi (**Table S5**). Analyses were performed in R (v4.6.1); analysis scripts are available as an online supplement on GitHub (https://github.com/mepepin/HCM_CPX.Diastology).

## Results

### Patient characteristics

The baseline analytic cohort comprised 441 patients (**Figure 1**) with mean age of 49.1 ± 15.6 years, 66.4% male; and median BMI 27.5 kg/m^2^ (IQR = 24.3-31.1 kg/m^2^). Left ventricular ejection fraction was preserved (median = 64.9%; IQR = 60.4%-69.0%; n = 281), and a resting LVOT gradient ≥30 mm Hg was present in 24.9% (69/277) of patients in whom it was recorded, and 19.0% had apical-variant HCM; consistent with prior HCM clinical trials, patients with mid-cavitary gradients were classified as non-obstructive. Median peak V̇O_2_ was 81.3% of predicted (IQR = 67.1%-97.9%) with a median V̇E/V̇CO_2_ slope of 30.9 (IQR = 27.5-35.5). Relative to 337 non-HCM reference subjects, patients with HCM had a greater mean interventricular septal dimension in diastole (1.7/1.9 cm among non-obstructive/obstructive vs. 0.9 cm for control), higher average LAVi (35.5/41.3 vs. 23.2 mL/m^2^ for control), and lower average e’ (7.0/5.7 vs. 11.8 cm/s for control) (**Table 1**).

**Table 1.**
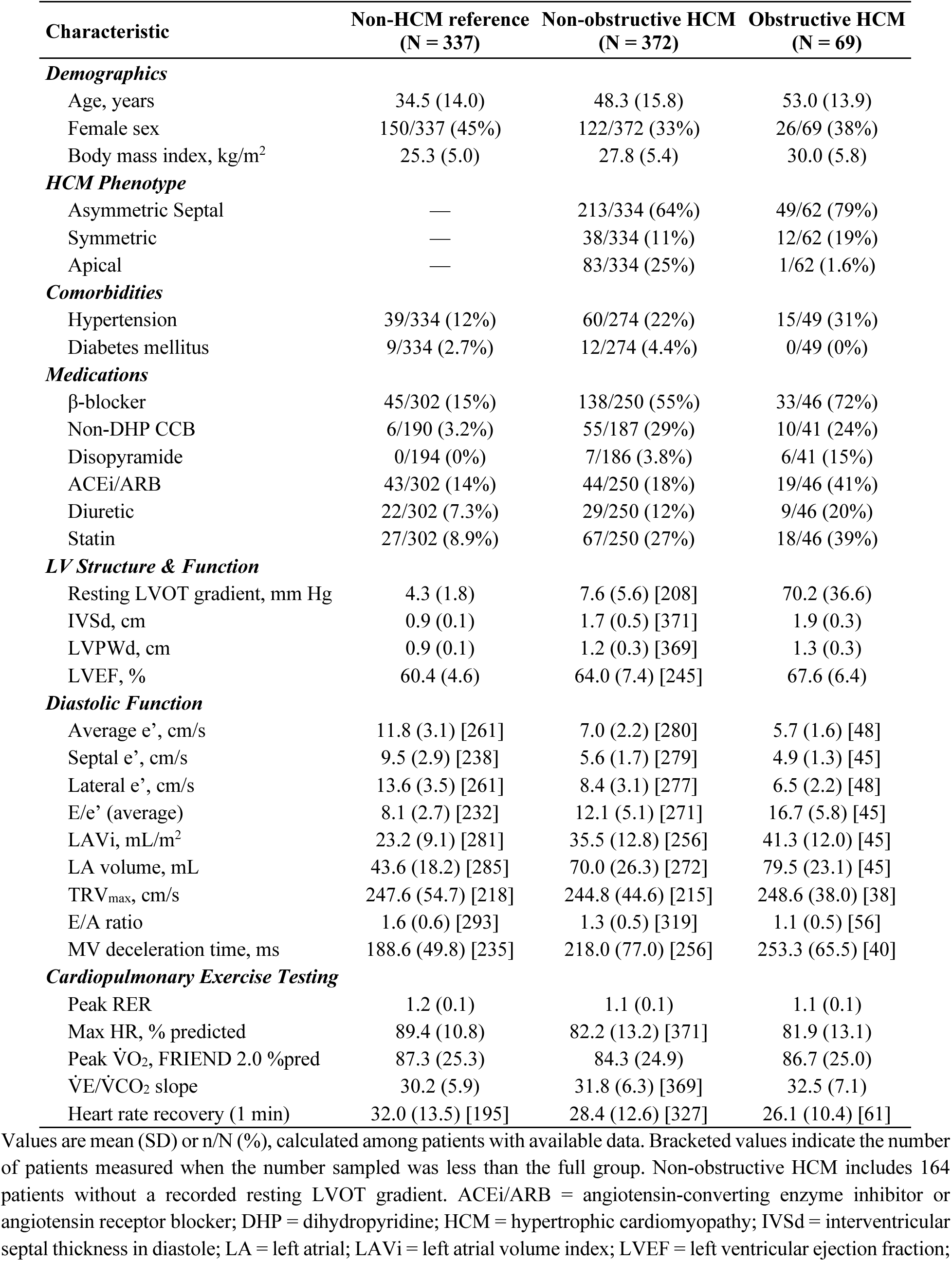
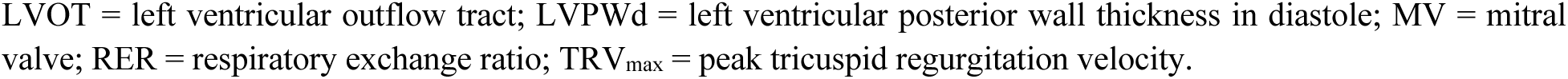
Baseline Characteristics of Patients With HCM and Non-HCM Reference Subjects.

| **Characteristic** | **Non-HCM reference**<br>**(N = 337)** | **Non-obstructive HCM**<br>**(N = 372)** | **Obstructive HCM**<br>**(N = 69)** |
| --- | --- | --- | --- |
| ***Demographics*** |  |  |  |
| Age, years | 34.5 (14.0) | 48.3 (15.8) | 53.0 (13.9) |
| Female sex | 150/337 (45%) | 122/372 (33%) | 26/69 (38%) |
| Body mass index, kg/m<sup>2</sup> | 25.3 (5.0) | 27.8 (5.4) | 30.0 (5.8) |
| ***HCM Phenotype*** |  |  |  |
| Asymmetric Septal | — | 213/334 (64%) | 49/62 (79%) |
| Symmetric | — | 38/334 (11%) | 12/62 (19%) |
| Apical | — | 83/334 (25%) | 1/62 (1.6%) |
| ***Comorbidities*** |  |  |  |
| Hypertension | 39/334 (12%) | 60/274 (22%) | 15/49 (31%) |
| Diabetes mellitus | 9/334 (2.7%) | 12/274 (4.4%) | 0/49 (0%) |
| ***Medications*** |  |  |  |
| β-blocker | 45/302 (15%) | 138/250 (55%) | 33/46 (72%) |
| Non-DHP CCB | 6/190 (3.2%) | 55/187 (29%) | 10/41 (24%) |
| Disopyramide | 0/194 (0%) | 7/186 (3.8%) | 6/41 (15%) |
| ACEi/ARB | 43/302 (14%) | 44/250 (18%) | 19/46 (41%) |
| Diuretic | 22/302 (7.3%) | 29/250 (12%) | 9/46 (20%) |
| Statin | 27/302 (8.9%) | 67/250 (27%) | 18/46 (39%) |
| ***LV Structure & Function*** |  |  |  |
| Resting LVOT gradient, mm Hg | 4.3 (1.8) | 7.6 (5.6) [208] | 70.2 (36.6) |
| IVSd, cm | 0.9 (0.1) | 1.7 (0.5) [371] | 1.9 (0.3) |
| LVPWd, cm | 0.9 (0.1) | 1.2 (0.3) [369] | 1.3 (0.3) |
| LVEF, % | 60.4 (4.6) | 64.0 (7.4) [245] | 67.6 (6.4) |
| ***Diastolic Function*** |  |  |  |
| Average e', cm/s | 11.8 (3.1) [261] | 7.0 (2.2) [280] | 5.7 (1.6) [48] |
| Septal e', cm/s | 9.5 (2.9) [238] | 5.6 (1.7) [279] | 4.9 (1.3) [45] |
| Lateral e', cm/s | 13.6 (3.5) [261] | 8.4 (3.1) [277] | 6.5 (2.2) [48] |
| E/e' (average) | 8.1 (2.7) [232] | 12.1 (5.1) [271] | 16.7 (5.8) [45] |
| LAVi, mL/m<sup>2</sup> | 23.2 (9.1) [281] | 35.5 (12.8) [256] | 41.3 (12.0) [45] |
| LA volume, mL | 43.6 (18.2) [285] | 70.0 (26.3) [272] | 79.5 (23.1) [45] |
| TRV<sub>max</sub>, cm/s | 247.6 (54.7) [218] | 244.8 (44.6) [215] | 248.6 (38.0) [38] |
| E/A ratio | 1.6 (0.6) [293] | 1.3 (0.5) [319] | 1.1 (0.5) [56] |
| MV deceleration time, ms | 188.6 (49.8) [235] | 218.0 (77.0) [256] | 253.3 (65.5) [40] |
| ***Cardiopulmonary Exercise Testing*** |  |  |  |
| Peak RER | 1.2 (0.1) | 1.1 (0.1) | 1.1 (0.1) |
| Max HR, % predicted | 89.4 (10.8) | 82.2 (13.2) [371] | 81.9 (13.1) |
| Peak $\dot{V}O_2$, FRIEND 2.0 %pred | 87.3 (25.3) | 84.3 (24.9) | 86.7 (25.0) |
| $\dot{V}E/\dot{V}CO_2$ slope | 30.2 (5.9) | 31.8 (6.3) [369] | 32.5 (7.1) |
| Heart rate recovery (1 min) | 32.0 (13.5) [195] | 28.4 (12.6) [327] | 26.1 (10.4) [61] |

Values are mean (SD) or n/N (%), calculated among patients with available data. Bracketed values indicate the number of patients measured when the number sampled was less than the full group. Non-obstructive HCM includes 164 patients without a recorded resting LVOT gradient. ACEi/ARB = angiotensin-converting enzyme inhibitor or angiotensin receptor blocker; DHP = dihydropyridine; HCM = hypertrophic cardiomyopathy; IVSd = interventricular septal thickness in diastole; LA = left atrial; LAVi = left atrial volume index; LVEF = left ventricular ejection fraction;
LVOT = left ventricular outflow tract; LVPWd = left ventricular posterior wall thickness in diastole; MV = mitral valve; RER = respiratory exchange ratio; $TRV_{max}$ = peak tricuspid regurgitation velocity.

### HCM-specific diastolic phenotypes

Guideline indices of LVDD are interpreted as a composite, yet the extent to which these identify the same patients in HCM is unclear. In our HCM population, LVDD abnormality thresholds were exceeded in 30.1% (95/316) E/e’ (>14), 47.5% (143/301) for LAVi (>34 mL/m^2^), and in 13.8% (35/253) for TRV_max_ (>2.8 m/s), whereas average e’ fell below the age-specific reference limit in 66.4% (211/318) (**Figure 2A**). Among HCM patients in whom both E/e’ and LAVi were measured, indices were both abnormal in 21.5% (56/261); 24.1% (63/261) had elevated LAVi alone, 9.6% (25/261) had elevated E/e’ alone, and 44.8% (117/261) had neither (**Figure 2B**). V̇E/V̇CO_2_ slope also differed across phenotypes (Kruskal-Wallis *P* = 0.001), at 33.6 (IQR = 29.7-37.0) in patients with both E/e’ and LAVi elevated versus 29.9 (IQR = 27.0-34.4) in those with neither, whereas percent-predicted peak V̇O_2_ did not differ significantly across the 4 phenotypes (Kruskal-Wallis *P* = 0.06) (**Figure 2C-D**).

**Figure 2:**
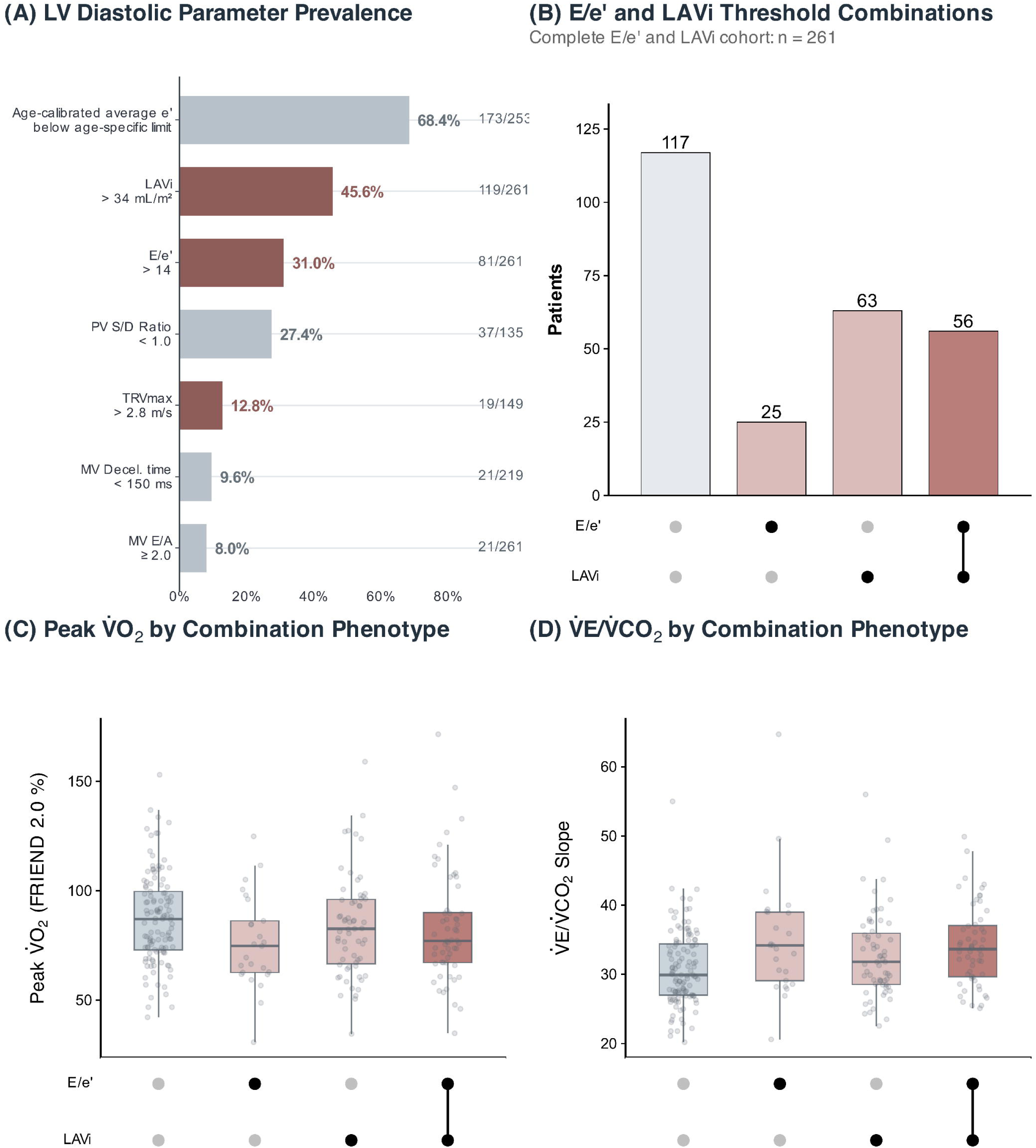
Diastolic phenotypes and cardiorespiratory fitness. **(A)** Prevalence of guideline-defined abnormalities of each diastolic index and of age-calibrated average e’. **(B)** Combinations of E/e’ >14 and LAVi >34 mL/m^2^ among the 261 patients with both indices measured. **(C)** Percent-predicted peak V̇O_2_ (FRIEND 2.0) and **(D)** V̇E/V̇CO_2_ slope across these combinations.

### Cross-sectional modeling of diastolic indices and fitness

Because guideline-based methods to characterize LVDD impose abnormality thresholds and are thus agnostic of severity, we modeled cardiorespiratory fitness using each diastolic parameter as a continuous variable (**Figure S2**). After adjustment for age, sex, and BMI, higher E/e’, LAVi, and TRV_max_ were each associated with a lower percent-predicted peak V̇O_2_ (partial F-test, all *P* ≤ 0.006), wherein effect sizes were similar across indices. Specifically, each SD increment corresponded to a lower peak V̇O_2_ of 6.0 (95% CI = 3.4-8.6), 3.6 (95% CI = 1.3-6.0), and 6.3 (95% CI = 3.5-9.2) percentage points of predicted for E/e’, LAVi, and TRV_max_, respectively (**Figure 3A-C**). Each index was likewise associated with a higher V̇E/V̇CO_2_ slope (E/e’, *P* = 0.045; LAVi, *P* = 0.002; TRV_max_, *P* < 0.001), with the largest effect for TRV_max_ at 2.5 units/SD (95% CI = 1.7-3.3) (**Figure 3D-F**). All 6 associations persisted after both FDR correction (q ≤ 0.045) and adjustment for septal thickness, resting LVOT gradient, hypertension, and beta-blocker or non-dihydropyridine calcium-channel blocker use (all *P* ≤ 0.045) (**Table S1**). Although most associations were approximately linear, models of E/e’ with peak V̇O_2_ and LAVi with V̇E/V̇CO_2_ slope exhibited nonlinear trends (nested F-test, *P* = 0.031 and *P* = 0.035, respectively). Specifically, the decline in peak V̇O_2_ attenuated beyond an E/e’ of 14.3 (*P* = 0.029), whereas V̇E/V̇CO_2_ slope rose at an inflection for LAVi (24.7 mL/m^2^, *P* = 0.020) and TRV_max_ (2.1 m/s, *P* = 0.049), both below their guideline thresholds and in a range where TRV_max_ is incompletely reported (**Table S6**).

**Figure 3:**
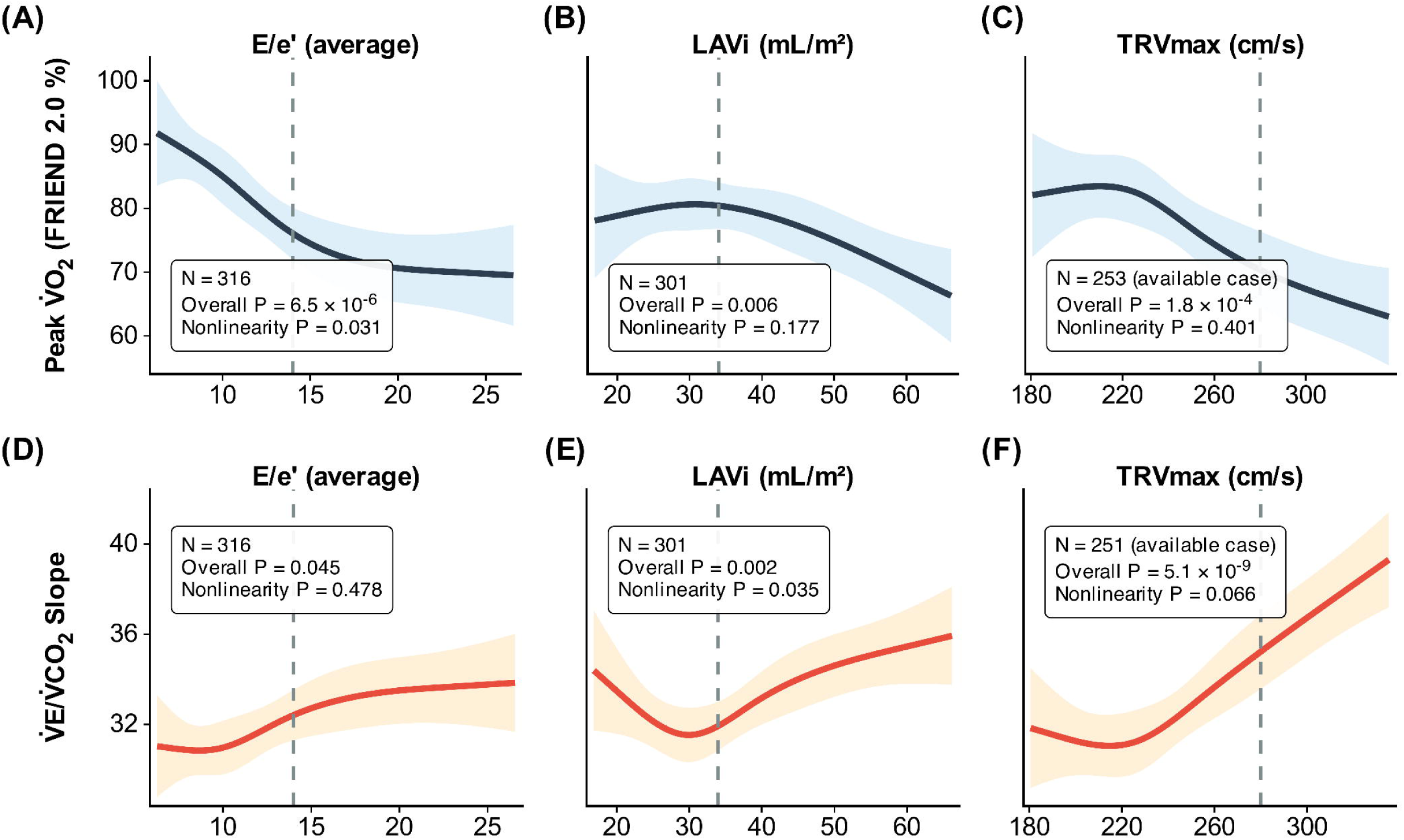
Cross-sectional associations of diastolic indices with cardiorespiratory fitness. Restricted cubic spline models, adjusted for age, sex, and BMI, relating E/e’ (n = 316), LAVi (n = 301), and TRV_max_ (n = 253) to **(A - C)** percent-predicted peak V̇O_2_ (FRIEND 2.0) and **(D - F)** V̇E/V̇CO_2_ slope (TRV_max_, n = 251). Shaded areas indicate 95% confidence intervals; dashed lines indicate guideline thresholds.

### Longitudinal association between diastolic dysfunction and cardiorespiratory fitness trajectory

To test whether baseline diastolic burden modifies the temporal trajectory of cardiorespiratory fitness, we modeled 615 CPETs obtained within 10 years of baseline in 199/441 patients with available repeat testing over a median of 3.1 years (IQR = 1.6-5.4 years; median 3 tests/patient); the number of patients contributing to each model varied with index availability (E/e’, n = 139; LAVi, n = 124; TRV_max_, n = 112 for peak V̇O_2_ and 111 for V̇E/V̇CO_2_ slope). In the primary mixed models, we found that E/e’, LAVi, and TRV_max_ did not significantly modify the trajectory of percent-predicted peak V̇O_2_ (interaction *P* ≥ 0.750) (**Figure 4A-C**), although generalized additive mixed models suggested nonlinear modification of peak V̇O_2_ trajectory by E/e’ (*P* = 0.022) and TRV_max_ (*P* = 0.002). In contrast, higher baseline LAVi was associated with a steeper rise in V̇E/V̇CO_2_ slope of 0.4 units/yr/SD (95% CI = 0.13-0.68; *P* = 0.005; q = 0.028; median follow-up 2.7 years in this model) (**Figure 4E**), whereas neither E/e’ (*P* = 0.777) nor TRV_max_ (*P* = 0.773) modified its trajectory. Therefore, LAVi emerged as the only diastolic index whose baseline level predicted a worsening ventilatory efficiency over time (**Figure S3**).

**Figure 4:**
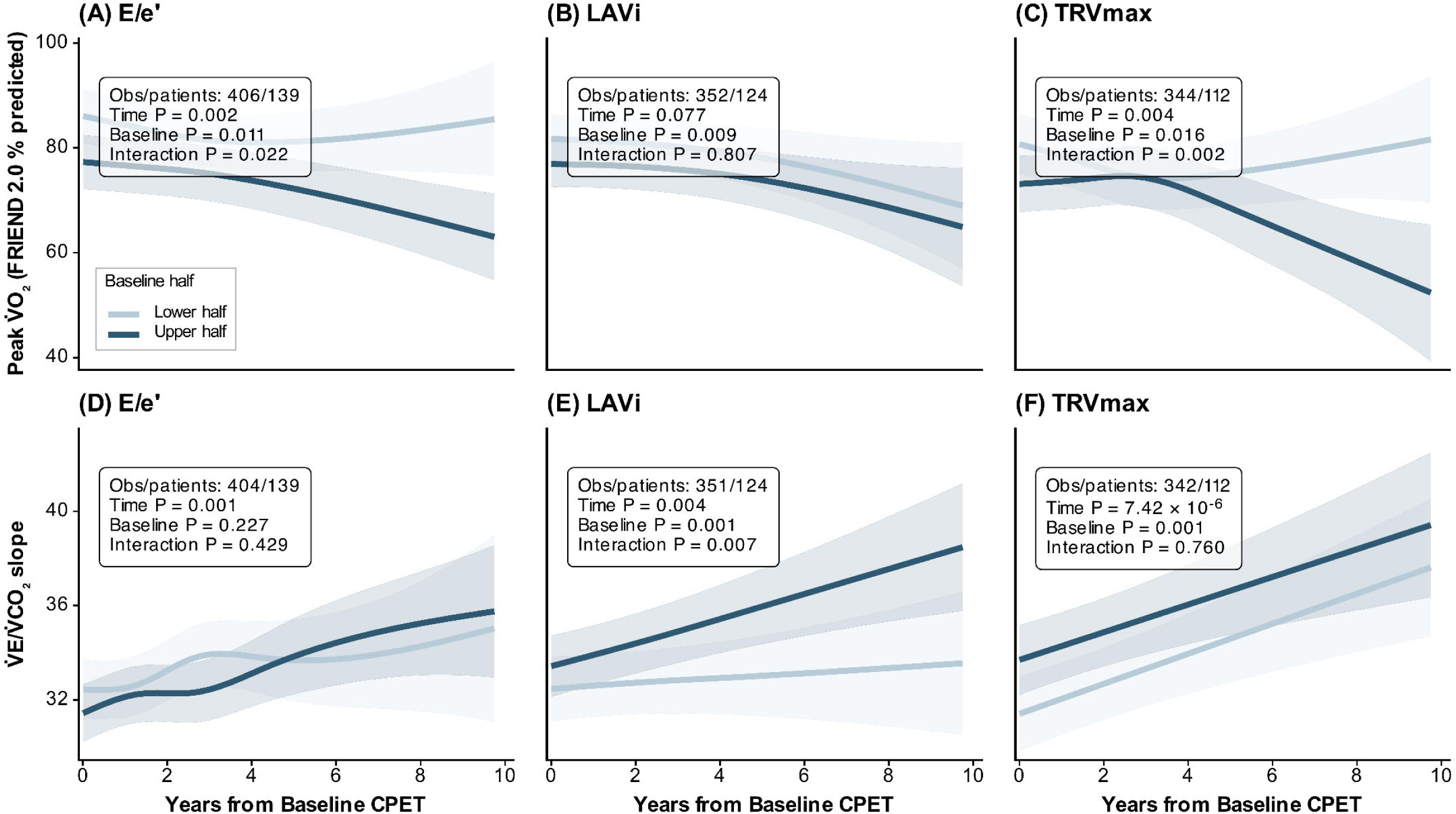
Longitudinal cardiopulmonary trajectories. Generalized additive mixed models (GAMMs) adjusted for age, sex, and BMI relating baseline E/e’, LAVi, and TRV_max_ to the subsequent trajectories of **(A - C)** percent-predicted peak V̇O_2_ (FRIEND 2.0) and **(D - F)** V̇E/V̇CO_2_ slope. Lines show predictions at the median values in the lower and upper halves of each baseline index; models used the continuous index. Inference on trajectory modification is from linear mixed models; curves show the corresponding generalized additive model predictions.

### Heart failure outcomes

To test whether LVDD identifies patients at higher risk for heart failure progression, we examined a sub-cohort of the study population for whom long-term clinical outcomes were reported. Among HCM patients followed for a median of 4.8 years (IQR = 1.5-8.0 years), 27.1% (118/436) reached the composite end-point at a median of 2.0 years (IQR = 0.5-4.7 years) after baseline CPET, of which 106 first events (89.8%) were hospitalizations for acute heart failure (**Table 2**). In Kaplan-Meier analysis, event-free survival differed across strata defined by E/e’ >14 (log-rank *P* = 0.001), LAVi >34 mL/m^2^ (*P* < 0.001), V̇E/V̇CO_2_ slope >30 (*P* = 0.006), and peak V̇O_2_ <80% of predicted (*P* < 0.001) (**Figure 5A-D**). In separate Cox models adjusted for age, sex, and BMI, each SD higher LAVi (HR = 1.68; 95% CI = 1.40-2.03; *P* < 0.001), E/e’ (HR = 1.36; 95% CI = 1.17-1.59; *P* < 0.001), and TRV_max_ (HR = 1.23; 95% CI = 1.02-1.47; *P* = 0.026) was associated with the composite end-point (**Figure 5E**). In parallel models, the strongest single association was with peak V̇O_2_ (HR = 1.74 per SD lower; 95% CI = 1.40-2.16; *P* < 0.001), followed by LV septal thickness (HR = 1.31; 95% CI = 1.09-1.59; *P* = 0.005) and V̇E/V̇CO_2_ slope (HR = 1.26; 95% CI = 1.07-1.48; *P* = 0.006). Proportional hazards held for every diastolic exposure (all *P* ≥ 0.220) (**Table S4**). These trends persisted with FDR correction and after adjustment for medications (**Table S3**, **Figure S4**).

**Figure 5:**
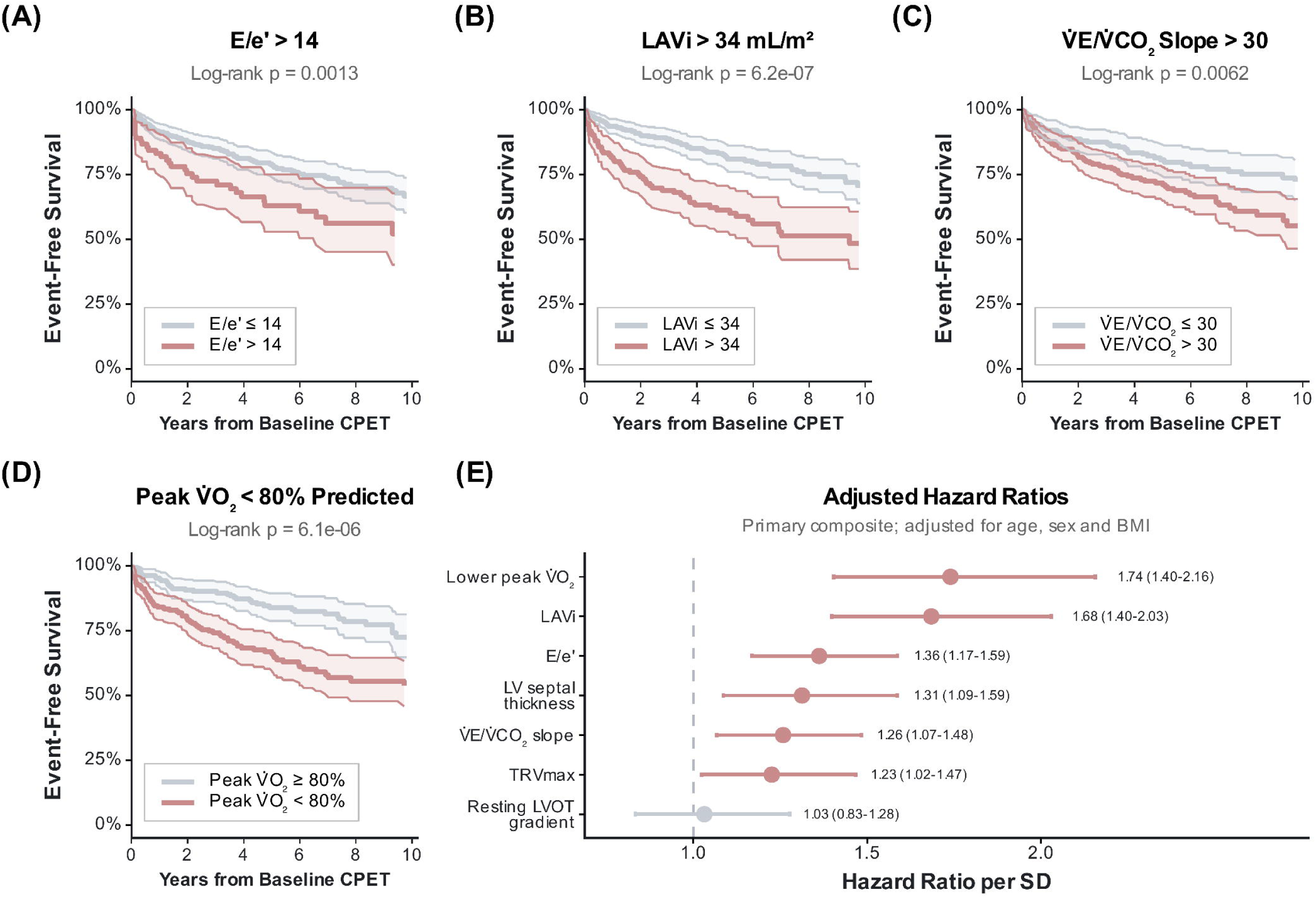
Association between diastolic indices and heart failure outcomes. Kaplan-Meier event-free survival according to **(A)** E/e’ >14, **(B)** LAVi >34 mL/m^2^, **(C)** V̇E/V̇CO_2_ slope >30, and **(D)** peak V̇O_2_ <80% of predicted (FRIEND 2.0). **(E)** Hazard ratios (95% CIs) per SD from Cox models adjusted for age, sex, and BMI; the peak V̇O_2_ hazard ratio is expressed per SD lower. Filled symbols denote *P* < 0.05.

**Table 2.** Composite Endpoint.

| **Event** | **Any occurrence, n (%)** | **First composite event, n (%)** |
| --- | --- | --- |
| Primary composite | 118 (27.1%) | 118 (27.1%) |
| Acute heart failure | 107 (24.5%) | 106 (24.3%) |
| Heart transplantation | 8 (1.8%) | 1 (0.2%) |
| All-cause death | 17 (3.9%) | 11 (2.5%) |
| Surgical myectomy (separate secondary outcome) | 51 (11.7%) | - |

Any-occurrence counts are not mutually exclusive. First-event counts identify the component that determined the time to the composite end point; components occurring on the same day were assigned to the most severe (death > transplantation > acute heart failure). Surgical myectomy was analyzed as a separate secondary end point.

## Discussion

Cardiopulmonary exercise testing is a cornerstone assessment for the risk stratification of HCM^26,27^ that offers the quantitative lens through which clinical efficacy is defined for HCM trials.^3,4^ Ventilatory efficiency carries prognostic weight across the heart failure spectrum,^28,29^ yet its relation to resting echocardiographic measures of LVDD, which are obtained in nearly every patient, has not been defined in HCM. In this single-center retrospective cohort, each resting diastolic index (E/e’, LAVi, and TRV_max_) was associated with both peak V̇O_2_ and V̇E/V̇CO_2_ slope, baseline LAVi was associated with subsequent worsening of ventilatory efficiency, and all 3 indices were associated with heart failure events, most strongly LAVi. These relationships were continuous across each index’s range and nonlinear for E/e’ with peak V̇O_2_ and for LAVi with V̇E/V̇CO_2_ slope. Together, these observations support a role for resting diastolic assessment in characterizing functional status and clinical trajectory in HCM.

In HCM, reduced peak V̇O_2_ is accompanied by impaired stroke volume augmentation that is most easily seen during peak exercise.^30^ The cardiac output-to-V̇O_2_ relationship becomes blunted, implicating a reduced stroke volume reserve rather than the impaired peripheral oxygen extraction typical of deconditioning.^31,32^ Resting Doppler indices are therefore unlikely to be direct determinants of cardiorespiratory reserve; instead, they likely reflect cumulative atrial and ventricular myocardial remodeling. This central relationship likely explains the comparable associations observed between each diastolic index and peak V̇O_2_. In fact, skeletal muscle and microvascular adaptations may counteract this cardiac constraint.^31,33^

The association we observed between resting diastolic indices and V̇E/V̇CO_2_ slope is physiologically intuitive. The V̇E/V̇CO_2_ slope reflects the ventilation required to eliminate a given quantity of CO_2_,^34^ and elevated pulmonary venous pressure raises it by compressing dependent capillary beds, thereby increasing alveolar dead space and producing ventilation-perfusion mismatch.^28,35^ TRV_max_ is the most proximate noninvasive estimate of pulmonary pressures, whereas LAVi and E/e’ may reflect structural and diastolic sequelae of chronically elevated left ventricular end-diastolic pressure.^36^ The relative strength of these associations may reflect the timescale each index integrates; TRV_max_ likely captures hemodynamic loading at the moment of imaging, whereas LAVi reflects remodeling that accrues over years, and E/e’ integrates myocardial compliance with preload. This distinction is consistent with the largest cross-sectional effect for TRV_max_ and the longitudinal signal confined to LAVi. In an earlier cohort of 156 patients with HCM from our institution, Finocchiaro et al. reported similarly that LAVi and E/e’ correlate with V̇E/V̇CO_2_ slope.^37^ Our findings therefore extend this observation to TRV_max_ and into a larger HCM cohort.

Baseline LAVi was associated with concurrent ventilatory inefficiency and its temporal decrement, whereas no index was significantly associated with the linear rate of change in aerobic capacity. In HFpEF, exercise-associated left ventricular filling pressures correlate with peak V̇O_2_,^38^ and in HCM ventilatory efficiency outperforms peak V̇O_2_ in detecting treatment effect.^39^ In secondary analyses of EXPLORER-HCM, for example, mavacamten improved ventilatory efficiency more than peak V̇O_2_.^3,40^ Because V̇E/V̇CO2 slope may more closely track hemodynamic change, a resting index that anticipates its deterioration, such as LAVi, may better identify patients for serial surveillance. By extension, left atrial enlargement could help identify candidates for earlier initiation of disease-modifying therapy, such as cardiac myosin inhibitors, though this warrants future testing.

The updated ASE framework was designed to both reduce the number of indeterminate cases seen in the 2016 algorithm^15^ and provide guidance for special populations including HCM^14^; however, it does not provide an evidence-based rubric to grade LVDD severity in HCM.^12^ Our data do not create severity grades; instead, each index correlates with fitness across its entire range, supporting that diastolic indices model fitness continuously across the HCM spectrum. LAVi more consistently outperformed E/e’. E/e’ is load dependent and geometrically constrained in HCM,^12,14^ whereas left atrial remodeling reflects cumulative diastolic burden and has been linked to exercise intolerance, ventilatory inefficiency, need for septal reduction therapy, and atrial fibrillation.^15,19,20,41^

### Limitations

Our observations should be interpreted within the context of a tertiary referral center with rapidly evolving HCM practice norms.^42^ The single-center design may have enriched for more complex phenotypes and limits generalizability to community practice, and the observational design supports association rather than prediction or causation. We did not assess Ar-A duration, provocative LVOT gradients, left atrial strain, or invasive hemodynamics. Diastolic indices were not universally measured, and availability differed modestly by outcome status (E/e’ less and TRV_max_ more often measured among patients with events), which may bias available-case estimates in either direction. Because left atrial enlargement predisposes to atrial fibrillation, which frequently precipitates heart failure hospitalization in HCM, incident atrial fibrillation may partly mediate the association between LAVi and heart failure-related events. Repeat CPET was clinically indicated rather than scheduled, so longitudinal analyses reflect the 45% of patients retested at variable intervals. Outcomes were ascertained from a single health system’s records, so events occurring elsewhere may have been missed. Finally, the cohort spanned nearly 2 decades and predated commercial availability of cardiac myosin inhibitors,^43^ whose effects on diastolic indices and prognosis warrant dedicated study.

## Conclusions

In this retrospective cohort of HCM, resting E/e’, LAVi, and TRV_max_ were each associated with aerobic capacity, ventilatory efficiency, and heart failure events. LAVi was the most informative, as it was associated with both subsequent deterioration of ventilatory efficiency and heart failure events, consistent with left atrial remodeling as a marker of cumulative diastolic burden. Interpreted as continuous measures rather than against dichotomous thresholds, resting diastolic indices may therefore add functional and prognostic information to an echocardiogram already obtained in nearly every patient with HCM, and LAVi may help identify patients who warrant closer clinical follow-up, serial exercise testing, and early consideration of disease-modifying therapy.

## Acknowledgments

The authors thank the Stanford University Hospital echocardiography and cardiopulmonary exercise testing laboratories for their outstanding clinical work.

## Data Availability

Analysis code is available at https://github.com/mepepin/HCM_CPX.Diastology; data are available from the corresponding authors upon reasonable request.

## Abbreviations and Acronyms

ASE: American Society of Echocardiography
CPET: cardiopulmonary exercise testing
HCM: hypertrophic cardiomyopathy
HFpEF: heart failure with preserved ejection fraction
LAVi: left atrial volume index
LVDD: left ventricular diastolic dysfunction
LVOT: left ventricular outflow tract
TRV_max_: peak tricuspid regurgitation velocity
V̇E/V̇CO_2_: minute ventilation to carbon dioxide production relationship
V̇O_2_: oxygen uptake

## Central Illustration

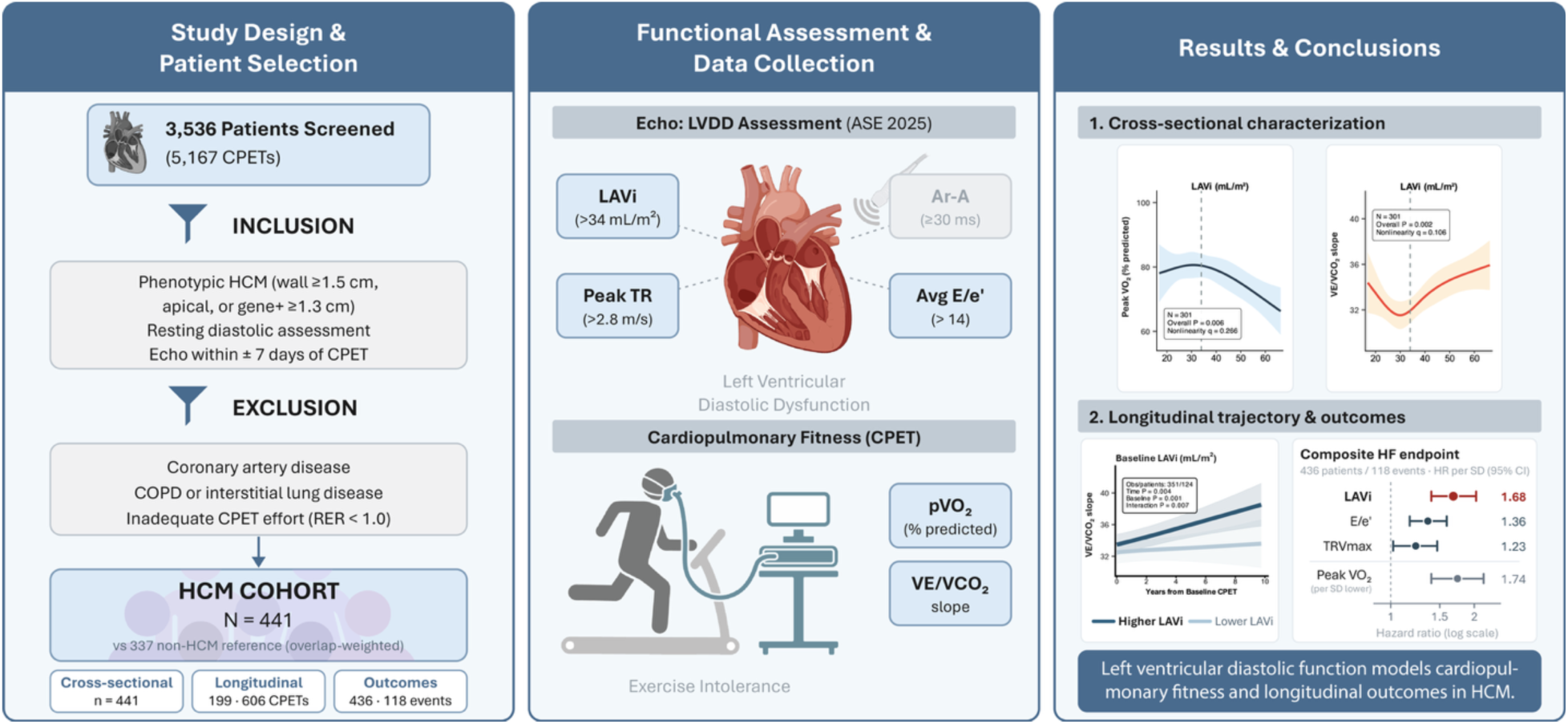

